# Investigation of mediating effects of sexual behaviours on the effect of a novel digital intervention on sexually transmitted reinfections: secondary analysis of a randomised controlled trial

**DOI:** 10.64898/2026.08.29.26361435

**Authors:** Isobel Landray, James R Carpenter, Caroline Free

## Abstract

**Background:** Preventing sexually transmitted re-infections brings health benefits and can be significantly less costly than treating their sequelae. Safetxt is a potential novel digital intervention developed to promote safer sexual behaviours. However, a recent randomised controlled trial of safetxt found no effect on reinfection at 1 year (OR 1.13, 95%CI: 0.98-1.31). We investigated if safetxt’s effect was mediated through sexually risky behaviours.

**Methods:** We used data from 6248 young people with STIs from 92 UK sexual health clinics. The direct and indirect effects of safetxt on reinfection were estimated using the counterfactual approach. Condom use at last sexual encounter, number of sexual partners and STI testing were assessed as mediators. These were analysed singly and together, using regression models and a formal weighting approach. The assumptions of each approach were considered and tested. Analyses were repeated in the subgroup showing the most promising effect of safetxt: men who have sex with men or with men and women (MSM/MSMW).

**Results:** No evidence was found for the total, indirect or direct effects differing from the null. Despite not being significant, for MSM/MSMW, some of safetxt’s effect on reducing reinfection was identified as being offset through its effect on number of sexual partners.

**Conclusions:** There was no evidence that safetxt’s effect on reinfection was mediated through changes in sexually risky behaviours. Adaptations to specifically target these behaviours are unlikely to improve safetxt’s overall effect. However, improving safetxt’s effect on the number of sexual partners a participant has may improve its effect for MSM/MSMW.

**What is already known on this topic:** The safetxt intervention did not result in a reduction in reinfection rate among 16-24 year olds diagnosed with chlamydia, gonorrhoea or non-specific urethritis in the UK. However, safetxt did result in increased condom use at 1 year. Results of the safetxt trial were most promising for men who have sex with men or with men and women (MSM/MSMW).

**What this study adds:** Disaggregating the effect of a novel digital intervention on sexually transmitted reinfections allows understanding of the mechanisms through which it acts. Investigators hypothesised that safetxt’s true effect was being mediated through changes in other sexual behaviours. This study shows there is no evidence of this from the safetxt trial.

**How this study might affect research, practice or policy:** All estimated effects were non-significant. Overall safetxt’s effect in the primary trial analysis was not due to mediation through condom use, number of sexual partners or STI testing. In MSM/MSMW, safetxt’s effect was identified as being reduced through number of sexual partners. Alterations to improve safetxt’s effect on the number of sexual partners a participant has could be considered with the aim of improving its overall effect on reinfection for MSM/MSMW.

## 1 Introduction

Sexually transmitted infections (STIs) are highly prevalent in young people and many are preventable through safer sex practices such as correct and consistent condom use [1–4]. Behavioural interventions are essential as no vaccines are currently licensed for most STIs [5–7].

One potential way to improve an individual’s sexual health and reduce their STI risk is through digital media. Young people are most at risk of STIs and use social media and messaging applications frequently [8, 9]. Therefore, a novel digital behavioural intervention ‘safetxt’ was developed with the aim to reduce STI incidence by increasing partner notification, condom use and STI testing before sex with new partners [10–14].

The safetxt trial assessed the effectiveness of the novel intervention delivered by text messages on sexually transmitted reinfections (STrI) in people aged 16-24 years in the UK [10]. The study was a parallel-group, individual-level, superiority, randomised controlled trial (RCT). Care providers and outcome assessors were blinded to allocation. A participant had to have been diagnosed or started treatment for chlamydia, gonorrhoea or non-specific urethritis (NSU) in the past two weeks. The trial randomised 6248 participants: 3123 to safetxt and 3125 to control. At 1 year, there were no benefits observed of safetxt on the primary outcome of reinfection (OR=1.13, 95%CI 0.98 to 1.31, p=0.08). Despite no benefit on the primary outcome, the qualitative feedback from safetxt participants was largely positive [12]. Safetxt recipients attributed increased condom use, STI testing after sex with new partners and confidence in partner notification to the intervention.

The primary result from the safetxt trial of no significant effect on risk of reinfection at 1 year was unexpected [10]. One proposed hypothesis to explain this finding is that safetxt’s effect on reinfection was mediated by sexually risky behaviours; receiving text messages designed to reduce stigma and blame during partner notification might have inadvertently resulted in increased number of sexual partners, which even if condom use increased, could increase risk of reinfection [2–4, 15]. It is of interest to work to disaggregate the null effect of the safetxt intervention into that which directly affected the reinfection rate, and that which was mediated by other behaviours throughout the study and assess if these effects worked in opposing directions [16]. Sexual health is especially prone to protective and risky behaviours working in opposing directions on overall STI infection rates.

This study aimed to assess the extent to which the effect of safetxt on reinfection was mediated in the whole study population, as well as in a subgroup that experienced the most promising effect of safetxt: men who have sex with men or with both men and women (MSM or MSMW) [10]. The analyses were extended to consider multiple mediators and how the mediators are inter-related.

## 2 Methods

### 2.1 Data source

The safetxt RCT provides individual level data for 6248 trial participants recruited from 92 sexual health clinics in the UK [10]. Participants were aged 16-24 years with a diagnosis of, or treatment for, chlamydia, gonorrhoea or NSU in the past two weeks and owned a mobile phone.

### 2.2 Safetxt intervention

Safetxt was a series of automated text messages to improve sex behaviours sent to participants over 1 year [10, 11, 17]. Safetxt’s aim was to reduce STI reinfection by increasing partner notification, condom use and STI testing before sex with a new partner. The message content and schedule were tailored by sex, sexual orientation and infection type at baseline.

Participants on the control arm received a monthly text for 1 year asking for any change in contact details.

### 2.3 Measures

Full details of trial measures assessed at all time points are reported in Suppl. File 1 [10, 11, 17]. Below, the measures relevant to this analysis are detailed.

Eleven variables measured at trial baseline were considered potential mediator-outcome confounders. The potential demographic confounders were: age, sexuality group, ethnicity, type of infection at baseline (chlamydia and/or gonorrhoea and/or NSU; unknown), education level (primary and secondary (age ≤16 years), secondary onwards (age ≥17 years), still in full time education) and index of multiple deprivation (IMD). Sexuality group was a variable created as specified in the trial Statistical Analysis Plan with groups as follows: MSM or MSMW; men who have sex with women only (MSW); women who have sex with men (WSM) or women who have sex with men and women (WSMW); women who have sex with women only (WSW); all other groups [18]. The potential yes-no sexual health confounders were: condom use during last sexual encounter, condom use during first sexual encounter with last new partner, tested before sex with last new partner, partner tested before sex with last new partner and number of partners in past 12 months (0, 1, ≥2). Any response of “unsure” in these self-reported variables were considered missing.

The mediators of interest were condom use at last sexual encounter and number of sexual partners (< or ≥ 2 partners) at 4 weeks and 1 year. The assessment of these measures are described elsewhere [10, 11, 17]. As part of an additional exploratory analysis, STI testing (from clinical records) at 1 year was also investigated as a mediator (Suppl. File 1).

The outcome measure was the same as for the primary trial analysis: reinfection at 1 year assessed by nucleic acid amplification tests [10].

### 2.4 Statistical analysis

The analyses were conducted with mediators treated as independent (“single mediators”) and as dependent (“multiple mediators”). The effect estimates are reported as risk differences.

#### 2.4.1 Single mediators

Each of the identified potential mediators were analysed separately. Because the outcome and all mediators considered were binary, the counterfactual approach could be utilised [19]. For each mediator under analysis, statistical models estimate the outcome each individual would be expected to have on each of the safetxt and control arm, with each of (i) the value of the mediator that is expected in the safetxt versus (ii) the value of the mediator expected in the control arm, conditional on their covariates. Having done this for each individual, averages could be taken to estimate the natural direct effect (NDE), natural indirect effect (NIE) and total treatment effect (TE). The STATA *mediate* command was used and the help file contains examples and details of the methodology [20].

#### 2.4.2 Multiple mediators

##### Composite mediators

A set of mediators were combined into a single composite variable [21] and then the “single mediator” analysis was conducted on this composite. This analysis was conducted with two composite mediators: condom use and sexual partners. If a participant used condoms at last sex at both 4 weeks and 1 year then their condom use composite was 1, and otherwise 0. If a participant had ≤1 sexual partner at both 4 weeks and 1 year then their sexual partners composite was 0, and otherwise 1.

##### Formal weighting approach

To estimate the NDE and NIE allowing for multiple mediators and treatment-mediator or mediator-mediator interactions, an inverse probability weighting (IPW)-based approach was implemented [22]. This formal strategy was conducted for two pairs of mediators: condom use at 4 weeks and 1 year, and number of sexual partners at 4 weeks and 1 year. Normal approximation and percentile-based bootstrap confidence intervals were estimated for the NDE, NIE and TE using 2000 bootstrap samples [23].

#### 2.4.3 Assumptions

To identify causal effects of interest, certain assumptions were made and considered carefully [10, 22, 24, 25–28]. Supplementary File 2 contains the details of the reasoning and tests of the validity of the assumptions of each approach.

## 3 Results

### 3.1 Single mediators

For all analyses of single mediators on all participants, the TE was positive: safetxt was associated with an increased risk of reinfection [10]. The NIE was negligible compared to the NDE (Table 1). Almost none of this effect of safetxt on reinfection acted through the mediator analysed. Within the MSM/MSMW subgroup, the TE was negative: safetxt was associated with decreased risk of reinfection. From the analyses of condom use at 1 year as a mediator, approximately 25% of the TE was through the mediator (−0.007/−0.029). The reinfection rate would decrease on average by 0.007 if everyone was on safetxt but the condom use at 1 year were changed from the level it would take if everyone was on the control arm versus on the safetxt arm (95%CI: −0.019 to 0.006). With the number of sexual partners at 1 year as a mediator, the direction of the NIE and NDE were opposite (mediation [29]). Safetxt was estimated to have a direct effect of reducing the rate of reinfection by 0.042 (95% CI: −0.131 to 0.046). However, the reinfection rate was estimated to increase by 0.014 if everyone was on safetxt but the number of sexual partners were changed from the level it would take if everyone was on the control arm versus on the safetxt arm (95%CI: −0.004 to 0.032).

**Table 1:**
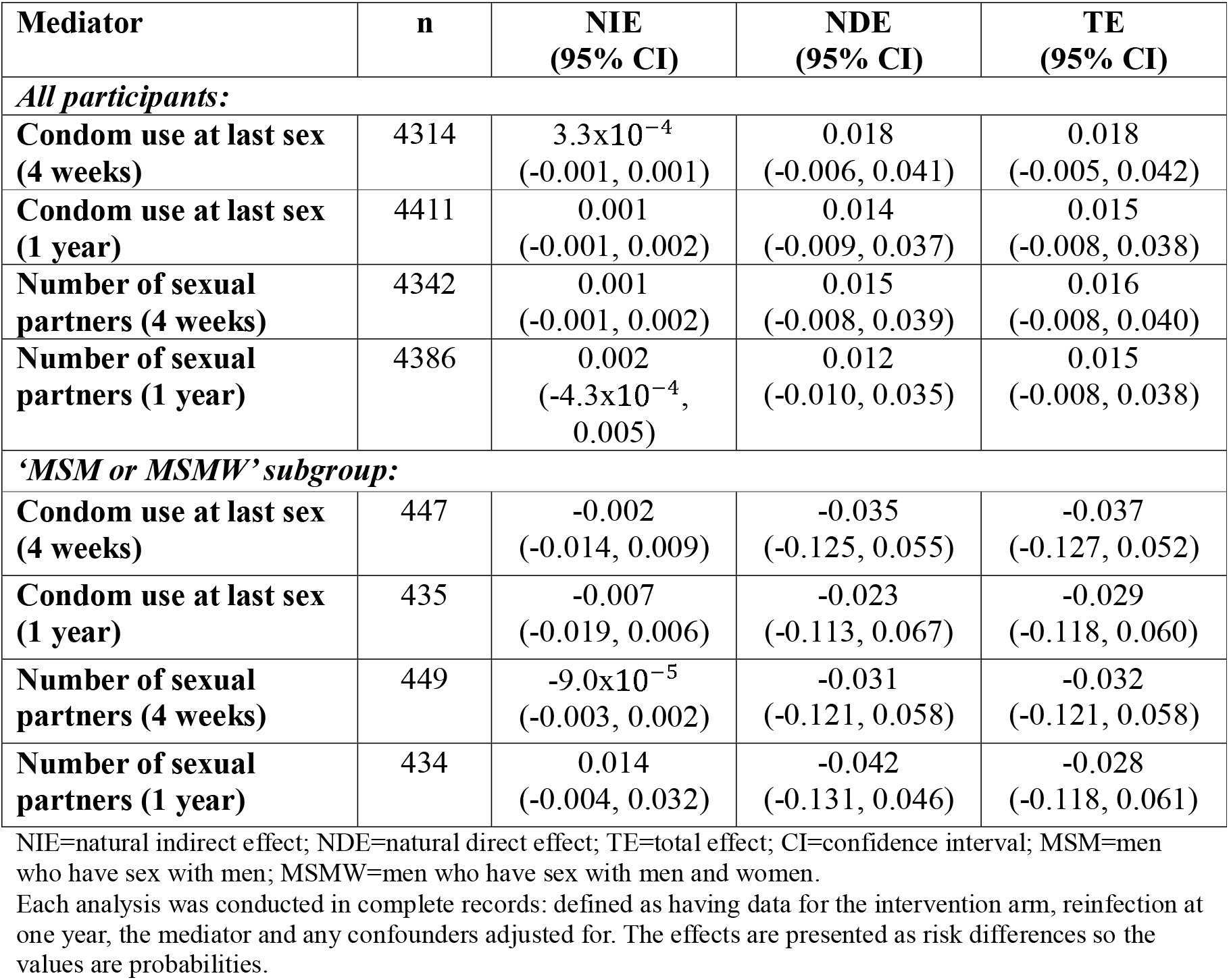
Risk difference estimates of intervention’s direct effect and indirect effect through mediators analysed independently on reinfection at one year.

### 3.2 Multiple mediators

#### Composite mediators

For both analyses in all participants, the NIE estimated through the composite mediator was very minimal (Table 2). The ‘harmful’ effect of safetxt on reinfection was not through the pathways with condom use or number of sexual partners. However, there were more substantial NIEs for MSM/MSMW. Approximately 15% of the protective TE of safetxt in the subgroup was through the condom use pathway (−0.004/−0.025). Some of the protective effect of safetxt on reinfection in this group may have been due to safetxt being associated with increased condom use which in turn reduces reinfection rates [10]. The direct protective effect of safetxt on reinfection in MSM/MSMW was offset by the number of sexual partners. Safetxt caused harm through the partners pathway; safetxt was associated with an increase in having multiple partners which in turn increases risk of reinfection [10]. The NIE through this composite mediator was approximately 20% of the magnitude of the NDE through all other pathways (0.010/0.047).

**Table 2:** Risk difference estimates of intervention’s direct effect and indirect effect through composite mediators on reinfection at one year.

| Composite mediator | n | NIE<br>(95% CI) | NDE<br>(95% CI) | TE<br>(95% CI) |
| --- | --- | --- | --- | --- |
| ***All participants:*** |  |  |  |  |
| **Condom use at last sex** | 4401 | $5.5 \times 10^{-5}$<br>($-3.0 \times 10^{-4}$,<br>$4.2 \times 10^{-4}$) | 0.017<br>(-0.006, 0.040) | 0.017<br>(-0.006, 0.040) |
| **Number of sexual partners** | 4308 | 0.003<br>($1.1 \times 10^{-4}$,<br>0.006) | 0.015<br>(-0.008, 0.038) | 0.019<br>(-0.005, 0.042) |
| ***'MSM or MSMW' subgroup:*** |  |  |  |  |
| **Condom use at last sex** | 438 | -0.004<br>(-0.014, 0.006) | -0.021<br>(-0.111, 0.069) | -0.025<br>(-0.115, 0.064) |
| **Number of sexual partners** | 446 | 0.010<br>(-0.010, 0.030) | -0.047<br>(-0.137, 0.043) | -0.037<br>(-0.126, 0.053) |

NIE=natural indirect effect; NDE=natural direct effect; TE=total effect; CI=confidence interval; MSM=men who have sex with men; MSMW=men who have sex with men and women.

#### Formal weighting approach

There were no extreme weights and there was fair overlap between the weights for each allocation arm (Table S2). For both analyses in all participants, the direction of the NIE was opposite to the NDE (Table 3). The magnitude of the NIEs and NDEs were very similar; the total effect of safetxt on reinfection was close to zero. When condom use at last sex at 4 weeks and 1 year were treated as mediators, safetxt’s effect on increasing condom use at last sex in turn reduced reinfection (−0.012, 95%CI: −0.036 to 0.012) while safetxt’s direct effect on reinfection was to increase its rate (0.014, 95%CI: −0.008 to 0.037). When the number of sexual partners at 4 weeks and 1 year were treated as mediators, safetxt’s effect on increasing the odds of having multiple partners in turn reduced reinfection (−0.010, 95%CI: −0.034 to 0.014) while safetxt’s direct effect on reinfection was to increase its probability (0.010, 95%CI: −0.013 to 0.034). There is substantial uncertainty in these results. but if the direction is correct they are counterintuitive. Having multiple sexual partners is generally associated with an increase in STI acquisition [15]. The direction of the NIEs and NDEs were reversed and closer to statistical significance in MSM/MSMW: safetxt increased reinfection through the condom use at last sex and number of sexual partners pathways.

**Table 3:**
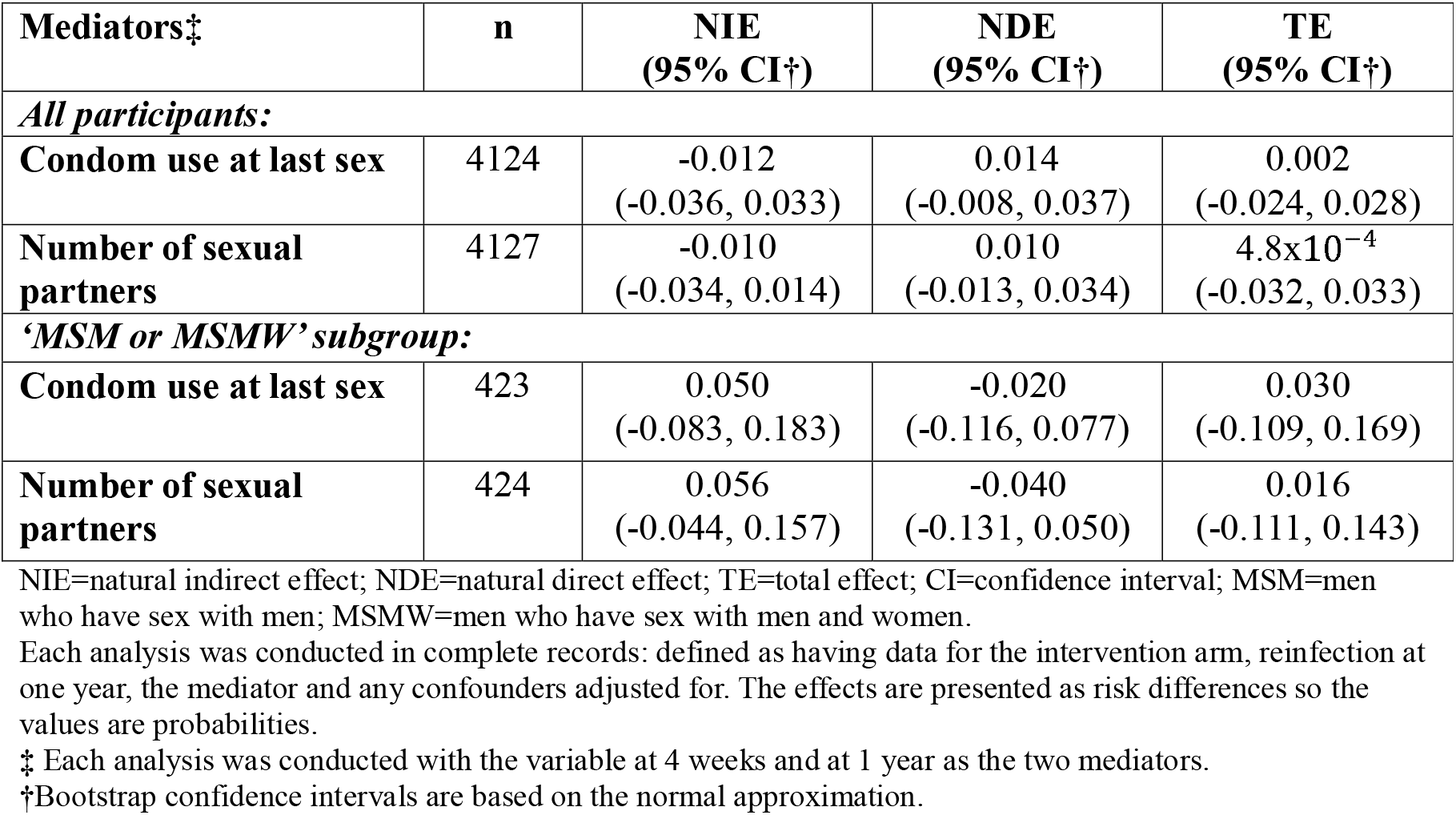
Risk difference estimates of intervention’s direct effect and indirect effect through multiple mediators on reinfection at one year.

In summary, none of the effects estimated from any of the analyses were significant at the 5% level and the CIs are wide (Figures S1 & S2).

## 4 Discussion

This mediation analysis aimed to improve understanding of the mechanisms through which the digital intervention ‘safetxt’ affects (or does not affect) the risk of STrI This allows for investigators to consider which causal pathways need targeting better for future interventions to be effective. Evaluation of suitable alterations to the safetxt strategy can therefore be discussed [30]. Sexual health education and prevention is an area of medicine where mediation analysis can be particularly informative. Policies and interventions can be more easily altered based on which causal pathways are, or are not, found to be affected.

When the potential mediators were analysed assuming no relationship between them, almost all of safetxt’s TE on reinfection was direct. However, for MSM/MSMW, some of safetxt’s effect on reducing reinfection was identified as being through its effect on condom use at last sex. The protective effect of safetxt for this subgroup was weakened by its effect on having multiple partners. The NDEs, NIEs and TEs from all three analyses (single, composite and multiple) were not significantly different from each other, or from no effect. From the effect point estimates, there is potentially some pattern that receiving safetxt was associated with higher odds of having multiple partners and this in turn was associated with higher odds of reinfection. While safetxt increased condom use in MSM/MSMW, the composite mediator and formal weighting approaches yielded estimates of opposing direction regarding safetxt’s indirect effect through condom use on reinfection. However, neither estimate was significant and the confidence intervals overlapped.

An analysis conducted with a similar goal to understand a sexual health intervention’s mechanism identified measures of self-efficacy to be strong mediators [31]. Further analyses could be conducted with similar mediators for safetxt.

### Strengths and limitations

Data used were from a large, representative sample of the population diagnosed with chlamydia and/or gonorrhoea [10]. Nonetheless, there were relatively low numbers of MSM/MSMW (n=593) limiting the subgroup analysis’s power. The follow up was high. Mediator-outcome confounders were identified based on a previous analysis with a pre-specified variable selection process and controlled for in the models [25].

The observed increase in number of sexual partners in the safetxt arm versus the control arm could have been due to social desirability bias.

The mediation analysis improved understanding of safetxt’s mechanism and effect on reinfection. A formal weighting approach was implemented to allow for analyses with multiple mediators [22]. The mediation analyses’ assumptions were assessed and deemed plausible.

All the analyses made an unlikely assumption that there is no relationship between the mediators ‘condom use at last sex’ and ‘number of sexual partners’ [27, 28]. The unbiasedness of effect estimates are contingent on the correct specification of the regression models fitted and the data being missing completely at random. However, based on the consistency of the results across analyses, we expect the null effects estimated to be robust.

## 5 Conclusions

Reactions to the lack of significant effect of safetxt on reinfection in the safetxt trial motivated mediation analyses to disaggregate the effect (or lack thereof) into that which acted through the two behaviours of interest (condom use and multiple partners), and that which was direct. The analyses suggested that very little of the total effect is mediated through condom use or number of sexual partners. Safetxt had most promising results in the MSM/MSMW so the mediation analyses were repeated in this group. Despite all estimated effects being non-significant, some of safetxt’s effect on reducing reinfection in this subgroup appeared to be acting through condom use and the effect was identified as being reduced through safetxt’s effect on the number of sexual partners. Alterations to improve safetxt’s effect on number of sexual partners a participant has could be considered with the aim of improving its overall effect on reinfection for MSM/MSMW.

## Acknowledgments

We would like to thank all those involved in the conduct of the Safetxt trial, including members of the trial steering committee, trial management group and team at LSHTM, collaborators and recruiting staff at all contributing trusts, and last, but not least all Safetxt participants. Isobel Landray would like to thank LSHTM where they were based when this work was carried out.

## Contributions of authors

CF was the Chief Investigator of the safetxt trial. CF contributed to the management of data collection. IL, CF and JRC conceived the idea for this secondary analysis. JRC was one of the trial statisticians, on whose SAP and code some of the analyses in this paper were based. IL conducted the analyses for this, with statistical supervision from JRC. All authors contributed to the interpretation of results. IL wrote the first draft of the manuscript with input from CF and JRC. All authors commented on revised versions and approved the final manuscript. CF is responsible for the overall content as guarantor and controlled the decision to publish.

## Ethics statement

We obtained ethics approval for the safetxt trial from the NHS Health Research Authority – London – Riverside Research Ethics Committee (REC reference 15/LO/1665) and the London School of Hygiene & Tropical Medicine (reference 10464). Participants provided informed consent in writing or via the trial website.

We obtained ethics approval for this analysis from the London School of Hygiene & Tropical Medicine (reference 30590).

## Data availability statement

Individual deidentified patient data, including a data dictionary, will be made available via our data sharing portal FreeBIRD website indefinitely. The trial protocol, statistical analysis plan, and trial publications will be available online. The Stata code for the analyses in this manuscript will be made available upon reasonable request.

## Patient and Public Involvement

No patient and public involvement was conducted for this secondary analysis.

## Supplementary materials

**Table S1:** Variables recorded in safetxt trial data.

Figure footnote: †*Each variables at this ‘timepoint’ is separated by ‘;’. The variables measured are statements or questions which participants answered or rated on appropriate scales e*.*g. from strongly disagree to strongly agree*.

**Table S2:** Summary of the weights applied to the participants in the multiple mediators analysis: the formal approach

Table footnote: *SD=standard deviation; MSM=men who have sex with men; MSMW=men who have sex with men and women;*

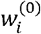 *and* 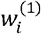 *are the weights applied in the formal weighting approach to handling multiple mediators*.

***‡*** *Each analysis was conducted with the variable at 4 weeks and at 1 year as the two mediators*.

**Supplementary file 1:** Additional analysis with clinical record of STI test at 1 year as mediator

**Supplementary file 2:** Assumptions of mediation analysis approaches

**Figure S1:** Risk difference estimates and 95% confidence intervals from each mediation analysis for all participants

Figure footnote: *CI=confidence interval (based on normal approximation). Some confidence intervals for the natural indirect effects are very narrow around the estimate*.

*Each analysis was conducted in complete records: defined as having data for the intervention arm, reinfection at one year, the mediator and any confounders adjusted for*.

*\*Condom use at last sex was a binary variable with value 1 only if used condoms at last sex at 4 weeks* ***and*** *at 1 year*.

*^Number of sexual partners was a binary variable with value 1 if the number of sexual partners was 2 or more at 4 weeks* ***or*** *at 1 year*.

*The effects are presented as risk differences so the values are probabilities. The grey dashed line indicates no effect (risk difference=0)*.

**Figure S2:** Risk difference estimates and 95% confidence intervals from each mediation analysis for ‘MSM or MSMW subgroup’

Figure footnote: *CI=confidence interval (based on normal approximation). Some confidence intervals for the natural indirect effects are very narrow around the estimate*.

*MSM=men who have sex with men only; MSMW=men who have sex with men and women*.

*Each analysis was conducted in complete records: defined as having data for the intervention arm, reinfection at one year, the mediator and any confounders adjusted for*.

*\*Condom use at last sex was a binary variable with value 1 only if used condoms at last sex at 4 weeks* ***and*** *at 1 year*.

*^Number of sexual partners was a binary variable with value 1 if the number of sexual partners was 2 or more at 4 weeks* ***or*** *at 1 year*.

*The effects are presented as risk differences so the values are probabilities. The grey dashed line indicates no effect (risk difference=0)*.

