## Supplementary figures and images for "Investigation of mediating effects of sexual behaviours on the effect of a novel digital intervention on sexually transmitted reinfections: secondary analysis of a randomised controlled trial"

### Figure S1

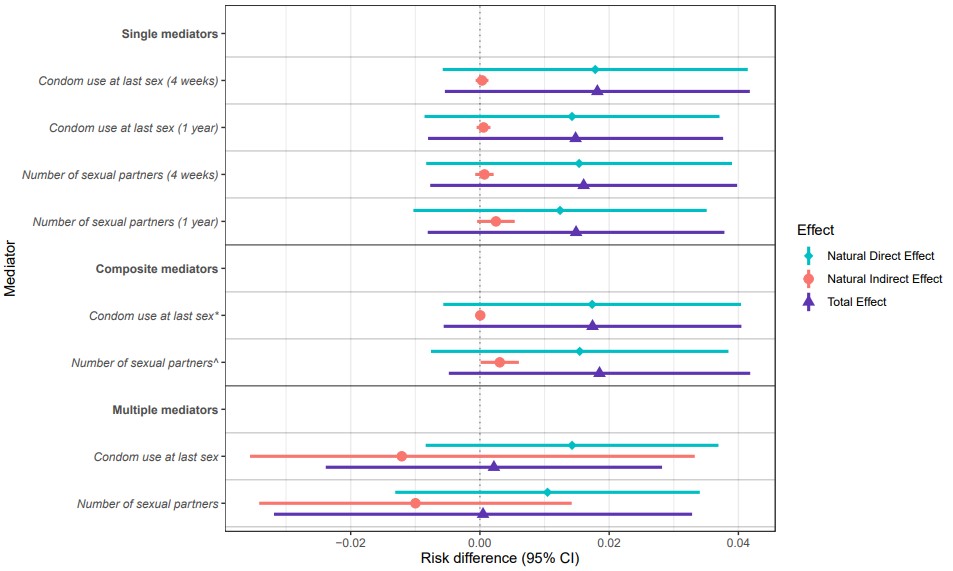

### Figure S2

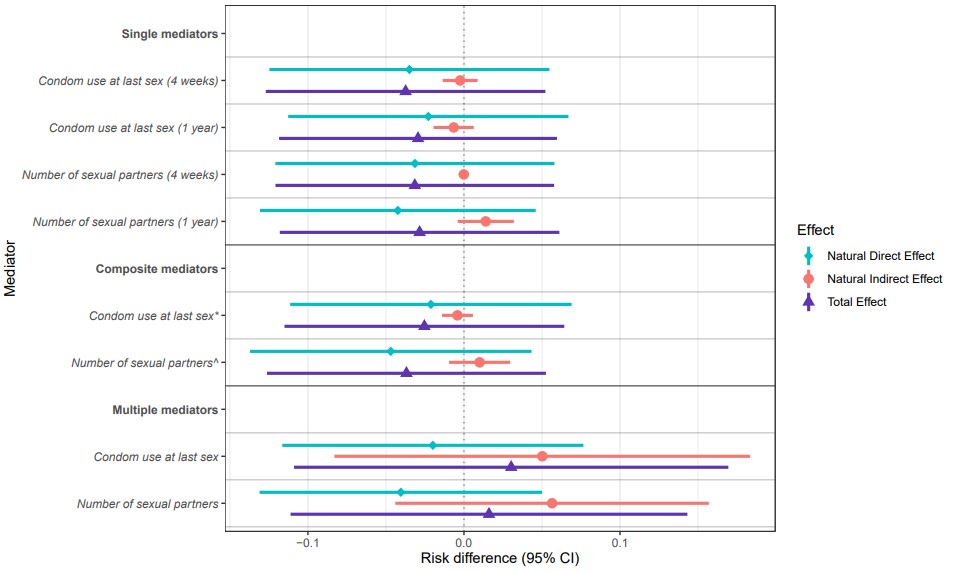
