## Supplementary file 1 for "Investigation of mediating effects of sexual behaviours on the effect of a novel digital intervention on sexually transmitted reinfections: secondary analysis of a randomised controlled trial"

**Supplementary File 1: Additional analysis with clinical record of STI test at one year as mediator**

Mediating effects of whether a participant had an STI test at one year on the effect of safetxt on reinfection at one year were assessed. The results are presented in Table S2.1. In all participants and the ‘MSM or MSMW’ subgroup, the total, indirect and direct effects were not significantly different from null effect. The magnitudes of the direct effects were larger than those of the indirect effects from the analyses with all participants and the 'MSM or MSMW’ subgroup only.

**Table: Risk difference estimates of intervention’s direct effect and indirect effect through STI testing behaviour analysed independently on reinfection at one year**

|  | **n** | **NIE**  **(95% CI)** | **NDE**  **(95% CI)** | **TE**  **(95% CI)** |
| --- | --- | --- | --- | --- |
| **All participants** | 4665 | 0.006 (-0.001, 0.013) | 0.014 (-0.008, 0.036) | 0.020 (-0.003, 0.043) |
| **‘MSM or MSMW’ subgroup** | 459 | -0.001 (-0.036, 0.035) | -0.041 (-0.122, 0.040) | -0.042 (-0.130, 0.046) |

NIE=natural indirect effect; NDE=natural direct effect; TE=total effect; CI=confidence interval; MSM=men who have sex with men; MSMW=men who have sex with men and women.

Each analysis was conducted in complete records: defined as having data for the intervention arm, reinfection at one year, the mediator and any confounders adjusted for. The effects are presented as risk differences so the values are probabilities.
