## Supplementary file 2 for "Investigation of mediating effects of sexual behaviours on the effect of a novel digital intervention on sexually transmitted reinfections: secondary analysis of a randomised controlled trial"

**Supplementary File 2: Assumptions of mediation analysis approaches**

The assumption of ‘conditional exchangeability’ in standard causal inference is extended in the mediation analysis setting. The analyses assumed there were; (1) no unmeasured treatment-outcome confounders given the covariates in the model, (2) no unmeasured mediator-outcome confounders given the covariates and the treatment, (3) no unmeasured treatment-mediator confounders given the covariates in the model, and (4) no unmeasured mediator-outcome confounders affected by the treatment given the covariates in the model. As safetxt allocation is randomised, (1) and (3) are automatically satisfied. However, randomisation does not guarantee control for mediator-outcome confounders and care needed to be taken to appropriately adjust for these. Previous secondary analyses involved model selection for variables predictive of the outcome (reinfection) and mediators [25]. For each analysis, covariates in common to at least one of the mediator models and the reinfection model were included to control for mediator-outcome confounding.

‘No interference’ is an assumption that the treatment arm of an individual does not affect the

potential outcomes of another individual. Safetxt messages encouraged partner notification and partner clinic attendance [10]. Exclusion of individuals known to be partners of participants already enrolled enables us to consider this assumption valid. Additionally, only 8.2% of participants reported that someone else read the messages they received.

‘Consistency’ is an assumption that for those who actually received safetxt/control, their observed outcome is the same as what it would have been had they received safetxt/control via the hypothetical intervention. Safetxt’s intended implementation is the same as in the trial.

In the ‘single mediators’ analyses, it was assumed that there was no mediator-mediator interaction influencing the outcome [26]. However, this working assumption was unlikely to hold fully. There were mediators that were repeated measures and were expected to be associated. The number of sexual partners a person has also influences their condom use at last sex [27, 28]. The ‘composite mediators’ analyses arbitrarily combine mediators. The assumption of no mediator-mediator interaction in the ‘multiple mediators’ analyses was only necessary between those chosen and those not.

For IPW in the ‘multiple mediators’ analysis to be valid, the probability of receiving safetxt and for having each level of the mediators had to be strictly between 0 and 1; both levels of the treatment and mediators were possible for all participants [22]. There also had to be some overlap in the weights when comparing treatment groups.

Both ‘single’ and ‘multiple’ mediator(s) analyses assumed that the models used to

estimate the causal effects were correctly specified [22].

As an exploratory analysis, all approaches were repeated in MSM/MSMW only. This was motivated by this subgroup showing the most promising effect of safetxt on reinfection.
