## Supplementary material for "Investigation of mediating effects of sexual behaviours on the effect of a novel digital intervention on sexually transmitted reinfections: secondary analysis of a randomised controlled trial": Table S1

**Table S1: Variables recorded in safetxt trial data, Free et al. (2022) [1]**

| **Timepoint** | **Variable type** | **Variables recorded** |
| --- | --- | --- |
| **Baseline** | ***Allocation*** | Intervention/control arm |
|  | ***Demographic*** | Age, sexuality*, gender, ethnicity, type of infection, education level, index of multiple deprivation. |
|  | ***Sexual health*** | Condom use during last sexual encounter, condom use during first sexual encounter with last new partner, tested before sex with last new partner, number of partners in past 12 months. |
|  | ***Contact details*** | Provision of: email address, alternative email address, alternative mobile number. |
| **Across the series of text messages†** | ***Knowledge related to STIs*** | If someone had an STI, they would know; STIs are rare; I can tell if someone has an STI |
|  | ***Attitude toward partner notification*** | Most people with STI will tell their partner; It is my responsibility to tell my partner if I had an STI; My partner would be glad I let them know I had an STI; My partner would think badly of me |
|  | ***Self-efficacy in telling partner about an infection*** | Ease to tell last partner you had STI; Ease to tell partner to get treatment; Ease to tell new partner you had STI; Ease to tell new partner to get treatment |
|  | ***Correct condom use self-efficacy*** | Ease to put condom on; Ease to stop condom drying out; Ease to stop condom breaking/coming off; Ease to keep condom on while withdrawing; Ease to keep condom on start to finish |
|  | ***Self-efficacy in negotiating condom use*** | Ease telling partner you want to use condoms; Ease telling new partner you want to use condoms; Ease telling new partner you won’t have sex unless use condoms |
|  | ***Reading intervention content*** | Did anyone else read the messages we sent you?; How did you feel about them reading the messages?; Did you know anyone else who took part in the study?; Did they read the messages we sent you?; Did you read the messages we sent you?; How many of the messages did you read? |
| **4 weeks follow-up** | ***Secondary outcomes*** | Correctly treated for STI; participant told last partner they had sex with before testing positive to get treatment; partner attended clinic for treatment; condom use at last sexual encounter |
|  | ***Other*** | Did you take the treatment for STI?; Did you avoid sex for 7 days after treatment?; Was a condom used last time you had sex?; Number of sexual partners since joining the study |
| **1 year follow-up** | ***Primary outcome*** | Cumulative incidence of chlamydia or gonorrhoea reinfection |
|  | ***Secondary outcomes*** | Condom use at last sexual encounter; ≥2 sexual partners since joining the trial; sex with someone new since joining the trial; condom use at first sex with most recent partner; STI testing for self, before first sexual encounter with most recent new partner (self-reported and testing confirmed by clinic record); most recent new partner was tested for STI before sex with participant; road traffic accident in past year when participant was driver; experience of partner violence in past year; diagnosis of “any” STI after joining trial according to postal test results and clinic records |

†Each variable at this timepoint is separated by ‘;’. The variables measured are statements or questions which participants answered or rated on appropriate scales e.g. from strongly disagree to strongly agree.

* Sexuality was grouped into the following: MSM or men-who-have-sex-with-men-and-women (MSMW); men-who-have-sex-with-women-only (MSW); women-who-have-sex-with-men (WSM) or women-who-have-sex-with-men-and-women (WSMW); women-who-have-sex-with-women-only (WSW); all other groups (non-binary individuals and individuals that did not state their sexuality were grouped due to data sparsity).

1. Free C, Palmer M J, McCarthy O L, et al. Effectiveness of a behavioural intervention delivered by text messages (safetxt) on sexually transmitted reinfection in people aged 16-24 years: randomised controlled trial. BMJ. 2022;378.
