## Supplementary material for "Investigation of mediating effects of sexual behaviours on the effect of a novel digital intervention on sexually transmitted reinfections: secondary analysis of a randomised controlled trial": Table S2

**Table S1: Summary of the weights applied to the participants in the multiple mediators analysis: the formal approach**

| **Mediators‡** | **n** | **Weight** | **Mean (SD)** | **Minimum** | **Maximum** |
| --- | --- | --- | --- | --- | --- |
| ***All participants:*** | | | | | |
| **Condom use at last sex** | 4124 | $w_{i}^{(0)}$ | 1.00 (0.04) | 0.86 | 1.32 |
|  |  | $w_{i}^{(1)}$ | 1.00 (0.05) | 0.80 | 1.21 |
| **Number of sexual partners** | 4127 | $w_{i}^{(0)}$ | 1.00 (0.04) | 0.93 | 1.27 |
|  |  | $w_{i}^{(1)}$ | 1.00 (0.03) | 0.82 | 1.09 |
| ***‘MSM or MSMW’ subgroup:*** | | | | | |
| **Condom use at last sex** | 423 | $w_{i}^{(0)}$ | 1.02 (0.15) | 0.66 | 1.21 |
|  |  | $w_{i}^{(1)}$ | 1.04 (0.24) | 0.84 | 2.38 |
| **Number of sexual partners** | 424 | $w_{i}^{(0)}$ | 1.02 (0.14) | 0.78 | 1.31 |
|  |  | $w_{i}^{(1)}$ | 1.03 (0.17) | 0.79 | 1.46 |

SD=standard deviation; MSM=men who have sex with men; MSMW=men who have sex with men and women;
$w_{i}^{(0)}$ and $w_{i}^{(1)}$ are the weights applied in the formal weighting approach to handling multiple mediators.

**‡** Each analysis was conducted with the variable at 4 weeks and at 1 year as the two mediators.
